# Trend and co-occurrence network study of symptoms through social media: an example of COVID-19

**DOI:** 10.1101/2022.09.28.22280462

**Authors:** Jiageng Wu, Lumin Wang, Yining Hua, Minghui Li, Li Zhou, David W Bates, Jie Yang

## Abstract

**Importance:** COVID-19 is a multi-organ disease with broad-spectrum manifestations. Clinical data-driven research can be difficult because many patients do not receive prompt diagnoses, treatment, and follow-up studies. Social media’s accessibility, promptness, and rich information provide an opportunity for large-scale and long-term analyses, enabling a comprehensive symptom investigation to complement clinical studies.

**Objective:** Present an efficient workflow to identify and study the characteristics and co-occurrences of COVID-19 symptoms using social media.

**Design, Setting, and Participants:** This retrospective cohort study analyzed 471,553,966 COVID-19-related tweets from February 1, 2020, to April 30, 2022. A comprehensive lexicon of symptoms was used to filter tweets through rule-based methods. 948,478 tweets with self-reported symptoms from 689,551 Twitter users were identified for analysis.

**Main Outcomes and Measures:** The overall trends of COVID-19 symptoms reported on Twitter were analyzed (separately by the Delta strain and the Omicron strain) using weekly new numbers, overall frequency, and temporal distribution of reported symptoms. A co-occurrence network was developed to investigate relationships between symptoms and affected organ systems.

**Results:** The weekly quantity of self-reported symptoms has a high consistency (0.8528, *P<0.0001*) and one-week leading trend (0. 8802, *P<0.0001*) with new infections in four countries. We grouped 201 common symptoms (mentioned ≥ 10 times) into 10 affected systems. The frequency of symptoms showed dynamic changes as the pandemic progressed, from typical respiratory symptoms in the early stage to more musculoskeletal and nervous symptoms at later stages. When comparing symptoms reported during the Delta strain versus the Omicron variant, significant changes were observed, with dropped odd ratios of coma (95%CI 0.55-0.49, *P<0.01*) and anosmia (95%CI, 0.6-0.56), and more pain in the throat (95%CI, 1.86-1.96) and concentration problems (95%CI, 1.58-1.70). The co-occurrence network characterizes relationships among symptoms and affected systems, both intra-systemic, such as cough and sneezing (respiratory), and inter-systemic, such as alopecia (integumentary) and impotence (reproductive).

**Conclusions and Relevance:** We found dynamic COVID-19 symptom evolution through self-reporting on social media and identified 201 symptoms from 10 affected systems. This demonstrates that social media’s prevalence trends and co-occurrence networks can efficiently identify and study public health problems, such as common symptoms during pandemics.

**Key points:** *Questions:* What are the epidemic characteristics and relationships of COVID-19 symptoms that have been extensively reported on social media?

*Findings:* This retrospective cohort study of 948,478 related tweets (February 2020 to April 2022) from 689,551 users identified 201 self-reported COVID-19 symptoms from 10 affected systems, mitigating the potential missing information in hospital-based epidemiologic studies due to many patients not being timely diagnosed and treated. Coma, anosmia, taste sense altered, and dyspnea were less common in participants infected during Omicron prevalence than in Delta. Symptoms that affect the same system have high co-occurrence. Frequent co-occurrences occurred between symptoms and systems corresponding to specific disease progressions, such as palpitations and dyspnea, alopecia and impotence.

*Meaning:* Trend and network analysis in social media can mine dynamic epidemic characteristics and relationships between symptoms in emergent pandemics.

## Introduction

The global coronavirus disease 2019 pandemic (COVID-19) caused by severe acute respiratory syndrome coronavirus 2 (SARS-CoV-2) has resulted in more than 616 million infections and 6.54 million deaths as of 28 September 2022.^1^ Furthermore, the pandemic is still ongoing, and its catastrophic impact may continue to grow and last for years. To broaden the understanding of this disease, relevant studies have been increasingly emerging, from determining molecular structures^2,3^ to developing drugs and vaccines.^4-6^ Concurrently, clinicians have endeavored to analyze clinical symptoms to guide therapeutic strategies.^7^ Public health officials have also tried to investigate the prevalence of symptoms to utilize the findings to provide precise prevention and control strategies for both people and governments.^8,9^

As a popular communication tool and public discussion platform, social media such as Twitter has permeated every aspect of our daily lives. Especially during the pandemic, social media played an essential role in information generation, dissemination, and consumption.^10,11^ There has been emerging COVID-19-related research based on social media. Such studies include topics in infodemics, public attitudes, detection or prediction of confirmed cases, and government responses to the pandemic^12^. However, they mainly focused on thematic analysis^13,14^ or sentiment analysis. ^15,16^ Only a few studies analyzed the symptoms and their epidemic-related characteristics.^17-19^ Moreover, these studies mainly conducted distribution and trend analyses in the early months of the pandemic rather than long-term, comprehensive investigations.

Current understandings of COVID-19 symptoms are primarily established on clinical data from medical institutions^20-22^, such as electronic health records (EHRs). However, nearly 80% of patients with asymptomatic or mild symptoms are not promptly or never clinically diagnosed and treated^23-25^, leading to potential missing information for mild and early symptoms. In addition, privacy policies on patient data have slowed cross-institutional cooperation and thorough studies of the pandemic on a large scale.^26^ Due to limited data sizes and sample diversity, current COVID-19 symptom network analyses only include a few typical symptoms, and do not construct a holistic network of comprehensive symptoms and affected systems.^27,28^

To attempt to address this research gap, we propose an efficient workflow for tracking and analyzing the general prevalence status and relationships of COVID-19 symptoms using social media.

## Methods

### Data collection

We selected non-retweeted English tweets related to COVID-19 using unique tweet identifiers from a widely used open-source COVID-19 tweet database.^29,30^ The tweets were identified by Twitter’s trending topics and selected keywords associated with COVID-19, such as *COVID-19* and *SARS-COV-2*. We downloaded 471,553,966 related tweets across 27 months, from February 1, 2020, to April 30, 2022, using Twitter’s Application Programming Interface (API).

### Symptom lexicon

Based on current literature, we built a comprehensive and hierarchical COVID-19 symptoms lexicon containing synonyms of symptoms and their affected body parts.^31-35^ Specifically, we appended colloquial variants frequently found on social media (**eMethod 1**). In addition, we grouped symptoms according to the affected organs and systems into 10 families^36,37^: cardiovascular, digestive, integumentary, musculoskeletal, nervous, reproductive, respiratory, urinary, sensory, and systemic. The final symptom lexicon contains 10 affected organs/systems, 257 symptoms, and 1808 synonyms (**Supplement Files 2**).

### Text preprocessing and rule-based filtering

To identify tweets with self-reported symptoms for subsequent analysis, we designed a three-step method that can be roughly summarized into filtering tweets with strict COVID-19 keywords, text cleaning, and matching of self-reported symptoms (**eMethod 2**). The overall workflow of this study is shown in **Figure 1**.

**Figure 1.**
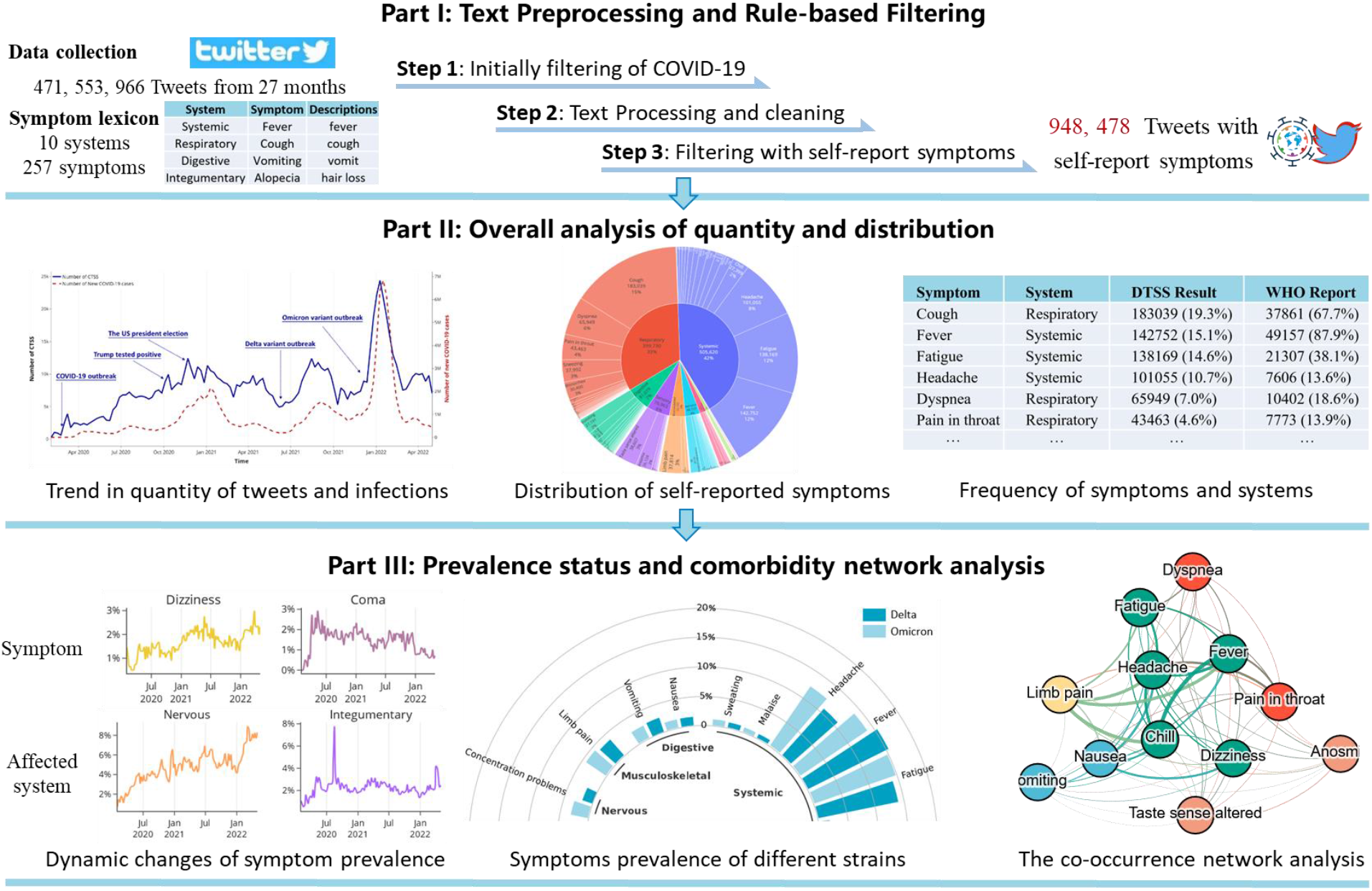
The overall workflow

### Trend analysis on the quantity of new COVID-19-related tweets

We compared weekly numbers of new COVID-19 tweets to new cases in countries with the most Twitter users. A survey on Statista shows that as of Jan 2021^38^, the top 4 countries with the most Twitter users and use English as their primary language are the United States (US), the United Kingdom (UK), the Philippines, and Canada (**eTable 2**). We used new COVID-19 cases in these countries reported by the World Health Organization (WHO) to be a rough representation of COVID-19 new cases (**Supplement Files 3**). We calculated weekly numbers of new tweets for both before and after the filtering. We also computed their Pearson correlation coefficient (*Pcc*) with the number of new cases to examine whether there was a statistically significant association between COVID-19 severity and public response.

### Overall distribution and dynamic frequency analysis of symptoms

Based on the COVID-19 symptom lexicon, we counted occurrences of each symptom by matching their synonyms against the filtered tweets datasets. Multiple mentions of the same symptom in one tweet were counted as one. To explore dynamic changes in symptom distribution with time, we calculated each symptom’s weekly frequency, normalized by the number of all self-reporting tweets. We also calculated the normalized frequency for each affected system.

### Comparison of symptoms prevalence status between different strains

COVID-19 has several variants that present different epidemic characteristics^39^, such as the highly transmissible B.1.617.2 (Delta) variant^40,41^ and B.1.1.529 (Omicron) variant^42^, which have led to rapid global rises. In this section, we compare self-reported symptom frequencies between Delta and Omicron. We extracted tweets from June 1, 2021, to Nov 27, 2021, when Delta was the globally dominant variant^36,43,44^ to represent Delta. Respectively, we extracted tweets from Dec 20, 2021, to Apr 30, 2022^36^ to represent Omicron.

We extracted symptoms from the two groups of tweets and selected those with ≥ 1% frequency as common symptoms. Then, we used the Chi-square test to calculate odds ratios (ORs) for Delta versus Omicron to assess the approximate prevalence differences of these common symptoms in two periods. Since a patient can get Delta in the Omicron-dominated period, this method calculates the odds of detecting a symptom among infected participants during the Delta-dominated period compared to the Omicron period.

### Network analysis

A COVID-19 patient may have multiple symptoms and report them simultaneously. Based on the symptom lexicon, we matched each symptom against each tweet to create a dataset **X** = [*x*_1_, *x*_2_, …, *x*_*n*_]∈ *R*^*n*×*m*^, where *x*_*i*_ = [*d*_*i*1_, *d*_*i*2_,…, *d*_*im*_]. *d*_*ij*_ is a binary feature that represents whether tweet *x*_*i*_ mentions symptom *j*; *m* and *n* represent the numbers of symptoms and tweets, respectively.

To quantitatively explore the strength of co-occurrence between two symptoms, we built symptom vectors **V**, where **V** = **X**^**T**^ =[*v*_1_, *v*_2_, …, *v*_*m*_]∈ *R*^*m*×*n*^, meaning that each dimension of *v*_*x*_ is a binary feature that indicates whether the symptom *x* was mentioned in tweet *i*. The co-occurrence strength is modeled by the similarity between the two symptom vectors, for which we adopted cosine similarity as the metric. In conclusion, the co-occurrence C between *v*_*x*_ and *v*_*y*_ can be modeled by the following equation:

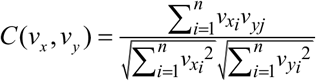

Based on the model, we constructed a weighted co-occurrence network of COVID-19 symptoms, where nodes represent symptoms and edges capture the co-occurrence strength between symptom pairs. We used Gephi^45^ and ForceAtlas2 algorithm^46^ to visualize the symptom network.

## Results

We selected 948,478 unique COVID-19-related tweets with self-reported symptoms to conduct these studies.

### Weekly trends of self-reporting tweets

We observed that weekly changes in self-reporting tweets were roughly consistent with the trends of new cases in the four selected countries (**Figure 2A**). The *Pcc* between the two trends is 0.8528 (*P*<0.0001), higher than the *Pcc* between new cases and the unfiltered COVID-19-related tweets (0.3235, *P*=0.0004, **eFigure 1)**. Moreover, self-reporting tweets showed a significant leading trend compared to the new cases when the leading time was set to one week. Such a trend had a higher correlation (*Pcc* = 0.8802, *P*<0.0001) than when no time difference was set.

**Figure 2.**
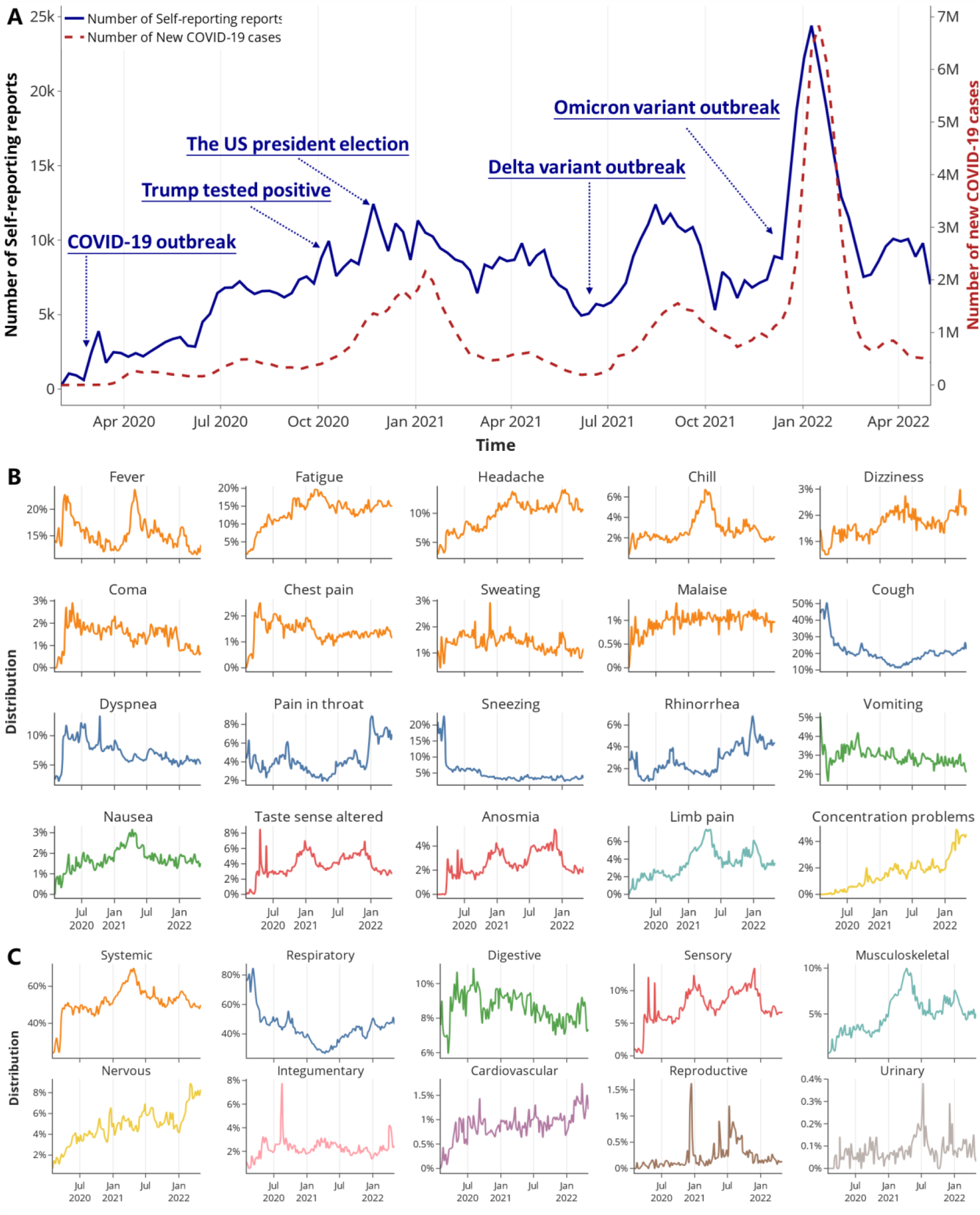
Weekly numbers of self-reporting tweets and weekly trends of the frequency of symptoms and affected systems (A) Weekly numbers of self-reporting COVID-19 tweets and sum of new COVID-19 cases in the US, the UK, Canada, and the Philippines. There were several waves of new cases and self-reporting tweets, including the initial outbreak in March 2020 and the continuous rapid spread. The first peak occurred during the transition of 2020 and 2021. Weekly new cases fell back to a pre-peak level and then increased at a slow rate until the outbreak of Delta, which started a new wave of infections in middle 2021. Omicron swept across countries from December 2021, took over Delta, and gave rise to the most enormous COVID wave. During the week of January 16, 2022, weekly new cases reached the highest number of 6.83 million. The weekly self-reporting showed similar trends but with more fluctuations. Such fluctuations mainly happened with hotspot issues on social media. One example was when former US president Donald Trump tested positive for COVID during the presidential election. (B) Weekly trends of the frequency of the top 20 symptoms and (C) Weekly trends of the frequency of the affected systems. Colors of symptoms in (B) correspond to affected systems in (C).

### Distribution of COVID-19 symptoms and affected organs/systems

In all, 245 symptoms were mentioned a total of 1197,733 times in 948,478 tweets. A total of 201 symptoms from 10 affected systems were mentioned in ≥10 tweets. The distribution of different systems and their related symptoms are hierarchically visualized in **eFigure 2**. Notably, systemic symptoms accounted for 42% of the total number of symptom occurrences, followed by respiratory (33%), digestive (7%), sensory (6%), musculoskeletal (4%), nervous (4%), integumentary (2%), cardiovascular (1%), reproductive (0.202%) and urinary (0.0645%) symptoms.

### Frequency of the common COVID-19 symptoms and affected systems

Overall, 20 common symptoms have more than a 1% frequency **(Table 1)** (more details in **Supplement Files 4)**. Note that the WHO report was based on 55,924 laboratory-confirmed cases from China in the early stage of COVID-19.^47^ The data of Delta and Omicron were extracted and calculated from our dataset in the corresponding period.

**Table 1.**
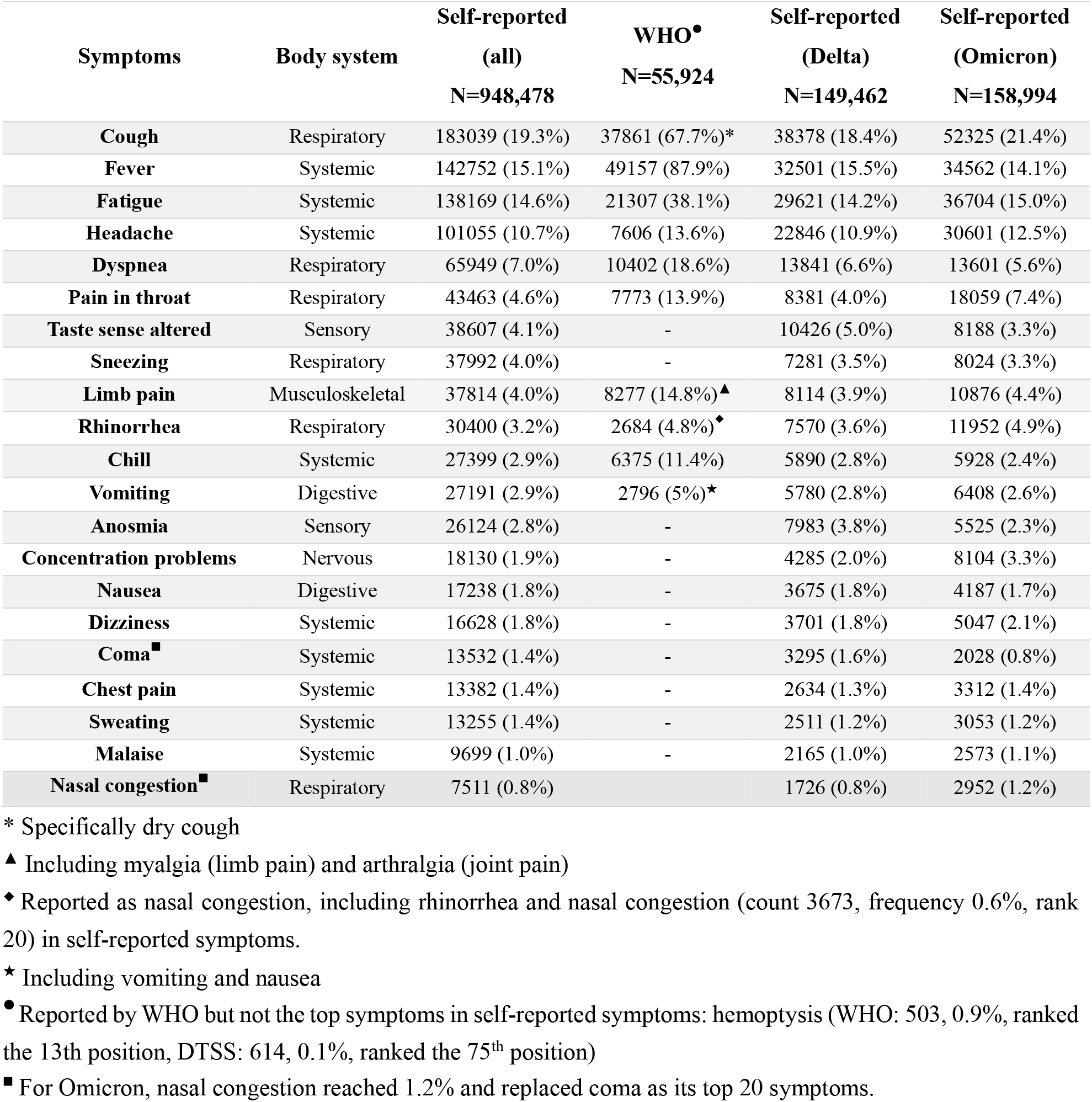
Occurrences and frequencies of common symptoms in filtered tweets

**Figure 2B** and **Figure 2C** show the weekly frequency of COVID symptoms and affected systems. The frequency of symptoms shows dynamic changes with the progression of the pandemic and has some distinct waves, respectively. In the early stage of COVID-19, cough, fever, and sneezing were the major symptoms, while other symptoms were rarely reported. With the progression of the pandemic, more symptoms, such as taste sense altered, chill, and anosmia, started to emerge. Respiratory symptoms were most common initially, accounting for more than 80% at one time, then gradually decreasing to about 40%. In contrast, the frequency of systemic, musculoskeletal, and nervous symptom mentions showed increasing trends. Frequencies of different symptoms gradually stabilized, with fluctuations associated with hotspot issues and the emergence of new variants.

### Distribution difference in symptoms between COVID-19 variants

The 209,074 tweets from June 1, 2021 to November 27, 2021 were placed in the Delta group, while 244,960 tweets from December 20, 2020 to April 30, 2021 were selected for the Omicron group. **Table 1** shows their top common symptoms and corresponding frequencies. **Figure 3** shows the ORs of common symptoms for Delta versus Omicron.

**Figure 3.**
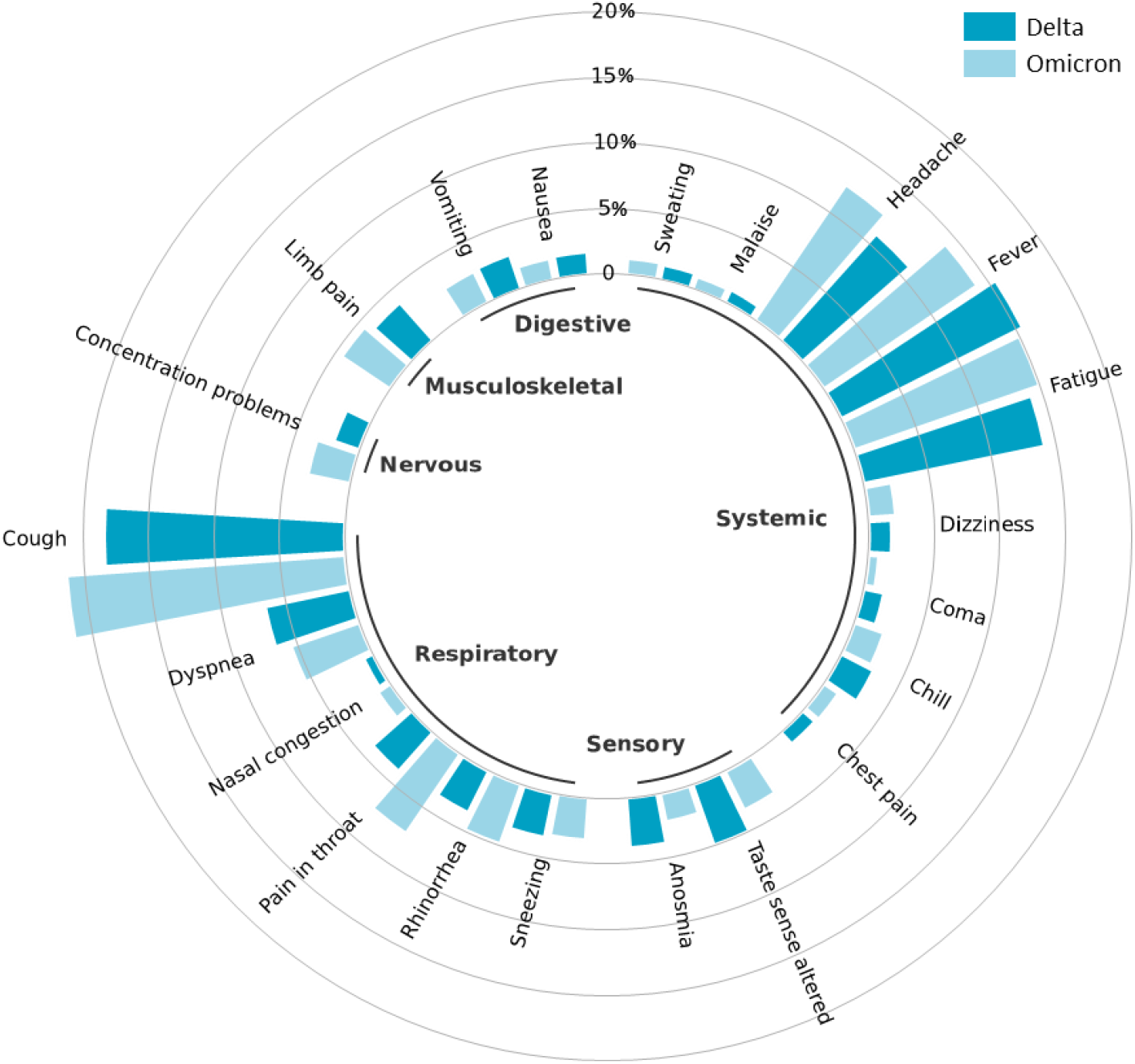
Distribution difference in common symptoms between Delta and Omicron

The top 20 symptoms of Omicron and Delta were roughly the same, but nasal congestion replaced coma as one of the top 20 symptoms of Omicron. Among these 21 symptoms, 8 were significantly (*P*<0.01) less prevalent (all P<0.01) among individuals infected during the Omicron period than Delta (top-5 OR: coma 0.52 [0.55-0.49], anosmia 0.58 [0.6-0.56], taste sense altered 0.66 [0.68-0.64], dyspnea 0.83 [0.85-0.81], chill 0.86 [0.89-0.82]), and 10 were significantly more likely to occur in Omicron patients than Delta (top-5 OR: pain in throat 1.91 [1.86-1.96], concentration problems 1.64 [1.58-1.70], nasal congestion 1.47 [1.38-1.55], rhinorrhea 1.37 [1.33-1.41], cough 1.21 [1.19-1.23])(More details in **eTable 3**).

### The co-occurrence network of COVID-19 symptoms

To simplify the co-occurrence network, we selected the top 100 symptoms by their overall distribution. The final network has 100 nodes with 2654 edges (**Figure 4**). Overall, the symptoms in this network show a clustering tendency according to the affected system, and the common symptoms are roughly distributed in the central region. Though systemic and musculoskeletal symptoms were not the leading part of the network, they are mainly in the center of the network and linked to the symptoms of different systems. Some outliers fall out of the clustering region of their theoretically affected systems. For example, palpitations, a cardiovascular symptom, locates at the center of the network next to systemic and musculoskeletal symptoms. Impotence, the only reproductive symptom with a high occurrence rate, and nocturnal enuresis, the only urinary symptom, located at the network border, demonstrating that co-occurrence with other symptoms were relatively low. Both intra- and inter-systemic symptoms had strong co-occurrences, such as chills and fever (both systemic symptoms), palpitations (cardiovascular) and dyspnea (respiratory), etc. For the readers to further explore the co-occurrences of a specific symptom, we provide an online visualization of the network (https://jgwu.top/COVID19-Symptoms-Twitter/network/).

**Figure 4.**
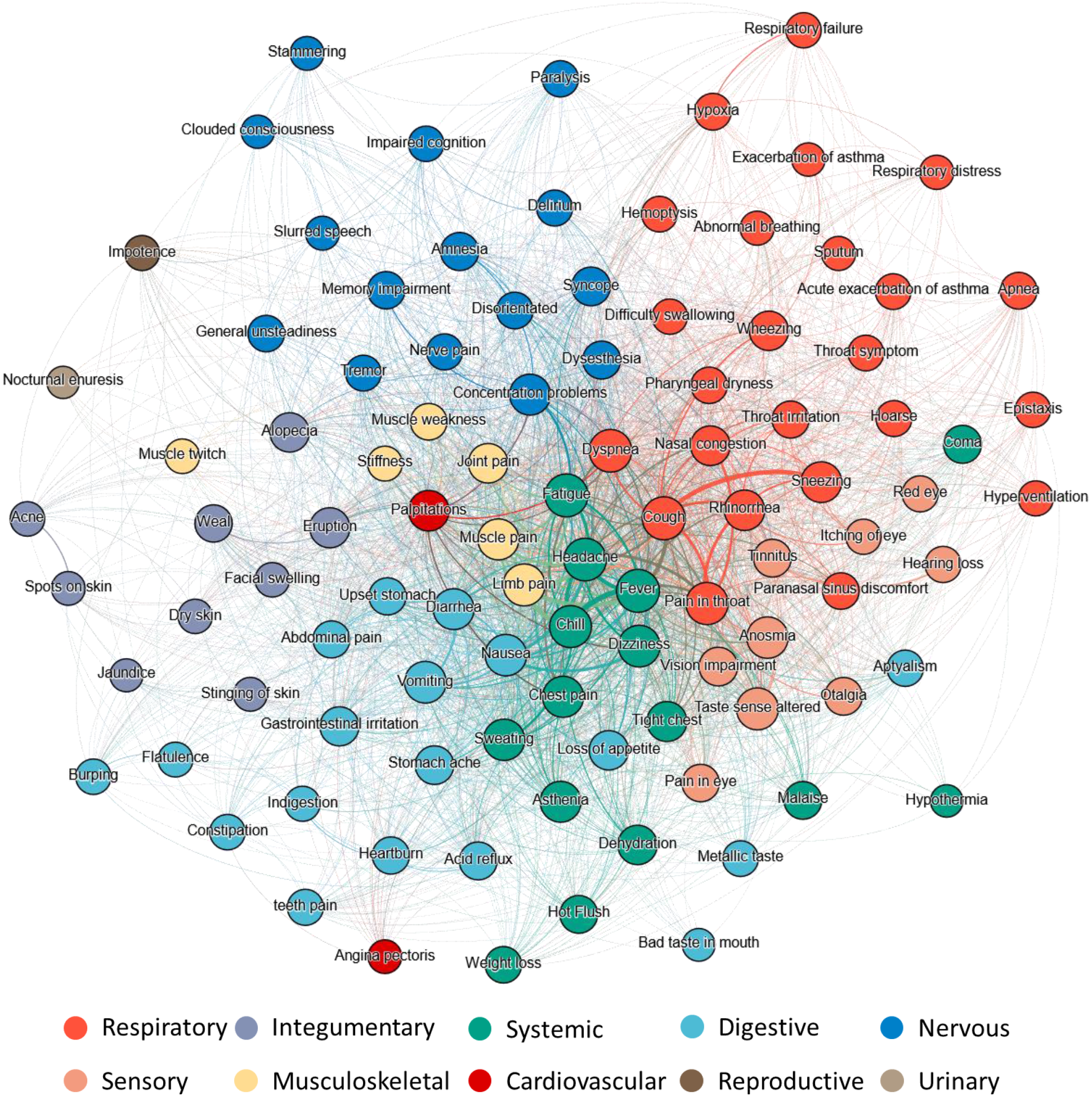
The co-occurrence network of different symptoms and affected systems.

## Discussion

In this work, we presented a novel workflow to investigate the symptom characteristics of an emergent pandemic using social media. We curated a hierarchical symptom lexicon that handles social media colloquialism and maps symptoms to their affected systems. This lexicon can be used in further social media-based medical research. We have also contributed a comprehensive co-occurrence network for COVID-19 symptoms for further exploration. To the best of our knowledge, this is the first dynamic prevalence status and network analysis of COVID-19 symptoms using large-scale and long-term social media data. This workflow can aid clinical professionals in monitoring unusual co-occurrent symptom patterns to promote pathogenesis studies. It is also promising in studying other emergent epidemics, given the accessibility and timeliness of social media.

Through the time trend analysis, we observed consistency between the trend of self-reporting tweets and new COVID cases (*Pcc*=0.8054). This suggests a highly positive significant correlation between the severity of the pandemic and the number of self-reported symptoms on social media. Masri et al. found that new case trends could be predicted one week ahead based on related tweets for the 2015 Zika epidemic.^48^ In correspondence and beyond, we found a highly correlated one-week leading trend of symptom-related tweets compared to new cases (*Pcc*=0.8802, *p*<0.0001) for COVID-19. This further demonstrates the sensitivity of social media and emphasizes the effectiveness of studying symptoms using social media in timely monitoring and prediction of pandemic status. Meanwhile, small fluctuations in the trends reflect public concerns with hotspot issues such as government policies and measures regarding the pandemic. For example, **Figure 2A** shows that the presidential election and Trump testing positive triggered increases in self-reporting tweets. This could be attributed to people discussing relevant problems and bringing up their own experiences, including symptoms. The insights gained from this type of trend analysis could help officials better guide and warn the public during pandemics. Readers can refer to our previous study for a more detailed investigation of the influence of hotspot issues on symptom reports.^49^

The common symptoms and their occurrence/prevalence ranks identified in our study are mostly in accordance with WHO reports but with different frequencies. These differences can be partially attributed to the different granularity and definition of symptoms. For example, cough in the WHO report only refers to dry cough, whereas wet cough is often correlated with sputum production.^47^ Such strict definitions are less suitable for self-reported social media data than traditional clinical studies. Using the adapted symptom lexicon, we identified a few symptoms that were not taken seriously in the WHO’s early reports, such as taste sense altered, anosmia, and nausea^50-52^. We also noticed some relatively infrequent symptoms, such as alopecia (occurrence: 5373) and impotence (occurrence: 2027). Recent studies have confirmed that SARS-CoV-2 may affect the expression of androgen and corrupt the physiological pathways involved in regulating erection.^53-55^ Having learned from the UK government’s experience of being urged by general practitioners to update the official COVID-19 symptom list to eliminate confusion^56,57^, policymakers should be aware that timely updates on the disease are essential to reassure the public, control the disease and better manage patients with specific complications.

**Figure 2** shows that symptom prevalence varied over time along with the virus evolution. As the key receptors of SARS-CoV-2 are highly co-expressive in the respiratory tract^58-60^, the initial symptoms are mainly respiratory and systemic symptoms caused by inflammation. However, over time, extensive self-reports of multiple symptoms from different systems confirmed that COVID-19 is a multi-organ disease^61^. At the later stage of the pandemic, there are increasing reports of persistent symptoms after COVID-19, such as fatigue, concentration problems, and limb pain (muscle/joint).^62,63^ Notably, consistent with recent findings on the increased risks of cardiovascular diseases^64^ and long neuropsychiatric symptoms^65^, our results show a burst of attention to nervous and cardiovascular symptoms on social media in January 2022, which have continued growing. This alerts us to the emerging prolonged signs (long-COVID)^66^ and their chronic burden on the nervous and cardiovascular systems.

Through the Chi-square test and **Figure 3**, we found that compared to Delta, as reported by the general population, Omicron has (1) lower ORs of severe symptoms, such as coma and dyspnea; (2) higher ORs for flu-like symptoms, such as pain in the throat, concentration problems, nasal congestion, and rhinorrhea; and (3) lower ORs of some typical COVID symptoms, such as anosmia and taste sense altered.^36,67,68^ This finding confirms that the Omicron is much more transmissible than previous variants but has less severe symptoms.^69,70^

The network of COVID symptoms and affected systems, built on massive data and a comprehensive lexicon, contains more extensive information than previous studies.^27,28^ While symptoms of the same system have higher co-occurrences, we did observe inter-system co-occurrences consistent with clinical studies. For example, coma exhibits strong relationships with respiratory symptoms in our networks, especially dyspnea, because the hypoxic/metabolic changes caused by intense inflammatory response trigger cytokine storm and may further result in coma and encephalopathy.^71^ We also found unusual co-occurrences. For example, palpitations as a cardiovascular symptom strongly correlate with dyspnea and dizziness (respiratory and systemic).^72^ Impotence, a reproductive symptom, has the strongest correlation with alopecia (an integumentary symptom), likely due to the SARS-CoV-2 invasion on androgen expression. They are also the typical long-COVID symptom among non-hospitalized patients.^73^ These strong relationships among unexpected group symptoms may point to new foci of disease progression or alert the potential risk of co-occurrent symptoms.

### Limitations

We acknowledge that our study has limitations. First, although we have reviewed substantial studies to construct a lexicon that is as comprehensive as possible, it inevitably misses some colloquial variants of the symptoms due to the noisy nature of Twitter. Second, the self-reported symptoms and cases are not laboratory-confirmed results. Moreover, some of our analyses could be biased. For example, we would expect more Delta patients in real life than in our study since people could still get Delta in the Omicron-dominated period. Therefore, we explicitly point out that our comparison is an estimation. Third, like every other public health study based on social media data, our study has potential cohort bias as the demographic distribution of social media does not represent that of the whole population.

## Conclusions

We developed a novel workflow to explore the dynamic characteristics of pandemic symptoms through social media. Using symptom analysis, we performed a large-scale and long-term social media-based study on COVID-19 and identified 201 symptoms from 10 systems. Compared to clinical data-based studies, we found a different symptom prevalence reported by a population of predominantly mild symptom patients. Evaluations like this can complement clinical studies to depict a more holistic picture of COVID-19 symptoms. The network reveals unusual co-occurrent symptom patterns, which may enable downstream pathogenesis studies. Thanks to the accessibility and timeliness of social media, this workflow is also promising in contributing to future public health studies, such as studying other emergent epidemics.

## Data Availability

All data produced in the present study are available upon reasonable request to the authors

## Data availability

The code of this study is available at: https://github.com/Dragon-Wu/COVID19-Symptoms-Twitter.

## Acknowledgments

This study has no funding support.

## Author Contributions

J.W, L.W, and M.L performed data analysis and drafted the manuscript. J.Y. designed the study. J.W, L.W, and M.L developed the symptom lexicon. Y.H prepared the data, helped draft and revised the manuscript. L.Z., J.Y., and D.W.B. provided critical reviews. All authors reviewed the manuscript. J.W. takes responsibility for the integrity of the work.

