## Supplementary material for "Trend and co-occurrence network study of symptoms through social media: an example of COVID-19": Supplment file 1

### Supplementary Content

**eMethod 1:** Design of symptom lexicon

**eMethod 2:** Pipeline of text preprocessing and rule-based filtering

**eFigure 1:** Weekly numbers of COVID-19-related tweets and new COVID-19 cases in the US, the UK, Canada, and the Philippines.

**eFigure 2:** The distribution of mentioned symptoms and their affected body systems.

**eTable 1:** Keywords and regular expressions for text preprocessing and rule-based filtering

**eTable 2**: Leading 20 countries that use Twitter as of January 2021

**eTable 3**: The different ORs of common systems between the original and delta

**eMethods 1:** Design of symptom lexicon

The primary sources include the standard symptom corpus compiled by Wang et al.[1], Goss et al.[2] based on the EHRs, the COVID-19 symptom corpus compiled by Sarker et al.[3], and the COVID-19 symptom keywords used in the study of Lopez-Leon et al.[4] and Mao et al.[5]. Since this study only focuses on physical and self-perceptible symptoms, we removed non-physical or non-perceptible symptoms and ambiguous abbreviations.

Since Twitter users often use personalized colloquialism rather than formal names to describe their symptoms, the same symptom can have many referents. As a result, we considered tense, person deixis, singular/plural forms, spelling mistakes, etc., for each symptom when curating the lexicon. We used the formal names defined in the SNOMED-CT (Systematized Nomenclature of Medicine Clinical Terms) [6] and added the varied forms of the proper names as their alternative names. For example, in our lexicon, the symptom "hearing loss" (proper name in SNOMED-CT) has descriptions (mostly from personalized colloquial descriptions) such as "deafness," difficulty hearing," and "loss of hearing," etc. We grouped symptoms according to their affected organs or systems into ten families, including cardiovascular, digestive, integumentary, musculoskeletal, nervous, reproductive, respiratory, urinary, sensory, and systemic. The final symptom lexicon contains 10 affected organs/systems, 257 symptoms, and 1808 descriptions.

**eMethods 2:** Pipeline of text preprocessing and rule-based filtering

(1) Initial filtering of COVID-19

The original keywords used to collect the COVID-19-related Tweets by Chen et al.[7] and Lopez et al.[8] were not strict enough, such as *China*, *Wuhan*, and *N95*. The tweets collected by these broad keywords were mainly about the pandemic outbreak rather than self-reporting about COVID-19. To improve the efficiency of subsequent analyses, we re-filtered the original dataset by matching tweets with at least one mention of keywords directly associated with COVID-19 (**eTable 1**). To filter negated tweets (e.g., "… don't have COVID-19 …"), we removed tweets containing negative indicators around COVID-19-related keywords. For example, tweets with *non-* before the keywords, and tweets with mentions of *no*, *ot*, *n't*, *nor,* etc. (**eTable 2**) within 5 words around the keywords.

(2) Processing and cleaning

To ensure that the tweets contain self-reported symptoms, we removed tweets with URLs since most of such tweets are retweets. To facilitate subsequent extraction of symptoms and analysis, we removed the non-English words and non-text contents, such as line breaks, extra spaces, emojis, and mentions of usernames. Notably, many phrases contain symptom-related keywords but do not mean symptoms, such as the symptom *tired* in the phrase *tired of* and the symptom *flush* in *flush the toilet.* Therefore, we applied regular expressions to mask such misleading patterns (**eTable 2**). Finally, we removed duplicated tweets based on the first 50 characters.

(3) Filtering with self-report symptoms

First-person-related keywords (i.e., I, I'm, me, my) and symptom descriptions from the lexicon were used to case-insensitively match against the filtered tweets.

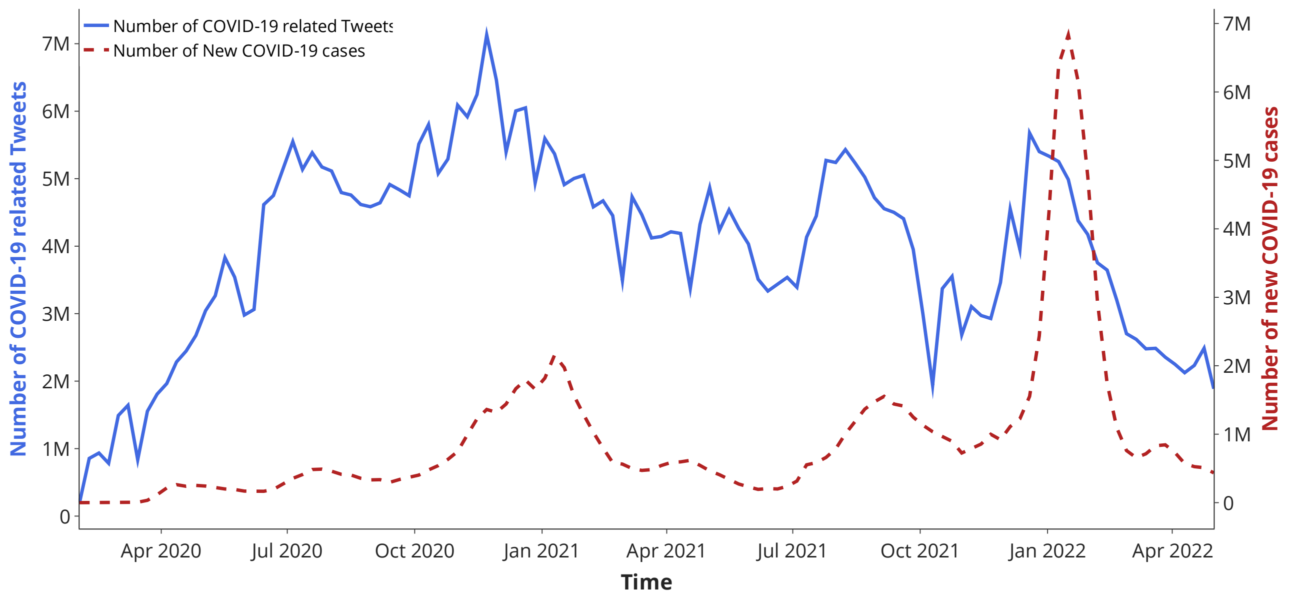


**eFigure 1.** Weekly numbers of COVID-19-related tweets and new COVID-19 cases in the US, the UK, Canada, and the Philippines.

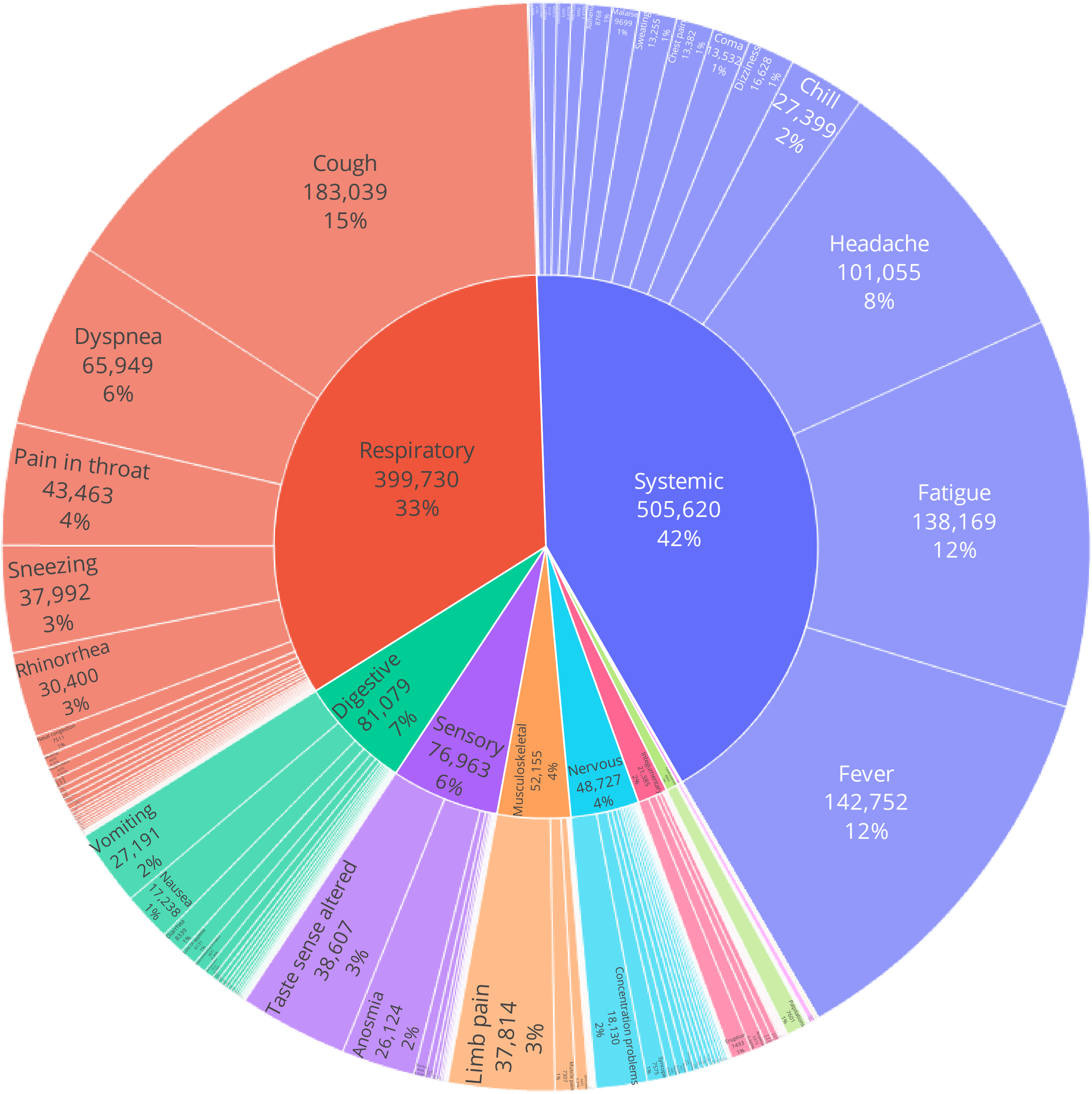


**eFigure 2.** The distribution of mentioned symptoms and their affected body systems.

**eTable 1.** Keywords and regular expressions for text preprocessing and rule-based filtering

<table>
<tr><th>Types</th><th>Examples</th></tr>
<tr><td>Strongly relevant to COVID-19</td><td>Coronavirus | corona virus | Koronavirus | Corona | Ncov | sars-cov-2 | sars cov 2 | sarscov | Wuhancoronavirus | chinese virus | chinesevirus | china virus | chinavirus | covid | Kungflu | kung flu | quarentinelife | stayhome | Epidemic | pandemic | pandemie | trumppandemic | trump pandemic | deltavariant | omicron | omicronvariant</td></tr>
<tr><td>Negative words</td><td>no | not | n't | n’t | never | none | neither | nor | without | lack | in the absence of | instead of | exclusive of | short of | rather than | hardly | scarcely | barely | seldom</td></tr>
<tr><td>Keywords of first-person</td><td>I | I’m | I am | me | my</td></tr>
<tr><td>Patterns for phrases that not mean symptoms</td><td>tired of | faint of | flush toilet | fake cough | in bad taste | a bad memory | (((cough(|ed|ing|s))|(sneez(e|ed|es|ing|y))) (cause|cuz|because|bc|in|over|on|works) ) | ((someone|always|could|every|fake|fakes|faked) ((cough(|ed|ing|s))|(sneez(e|ed|es|ing|y)))( |,|\.)) | ((tired )|((to|really|actually|already|so|don|look|this|are|were|'re|'s|was|is|your|our|her|his) tired(,|\.))) | ((a|the|is very|a bit|some|is|fake|faked|fakes) (faint(|ed|ing))( |,|\.))</td></tr>
</table>

**eTable 2.** Leading 20 countries that use Twitter as of January 2021

| Country | Twitter Users (in millions) | English is the Official Language | English is the Primary Language |
|---|---|---|---|
| United States | 69.3 | Yes | Yes |
| Japan | 20.9 | No | - |
| India | 17.5 | Yes | No |
| United Kingdom | 16.45 | Yes | Yes |
| Brazil | 16.2 | No | - |
| Indonesia | 14.05 | No | - |

| | | | |
|---|---|---|---|
| Turkey | 13.6 | No | - |
| Saudi Arabia | 12.45 | No | - |
| Mexico | 11 | No | - |
| France | 8 | No | - |
| Philippines | 7.85 | Yes | Yes |
| Spain | 7.5 | No | - |
| Thailand | 7.35 | No | - |
| Canada | 6.45 | Yes | Yes |
| Germany | 5.8 | No | - |
| South Korea | 5.15 | No | - |
| Argentina | 5 | No | - |
| Egypt | 3.7 | No | - |
| Colombia | 3.35 | No | - |
| Malaysia | 3.35 | Yes | No |
| Total | 254.95 | - | - |

**eTable 3.** The different ORs of common systems between the original and delta

| SNOMED | OR | P | 95% CI | Chi2 | Count and prevalence of Delta | Count and prevalence of Omicron |
|---|---|---|---|---|---|---|
| Coma | 0.5213 | <0.01 | [0.55 0.49] | 544.8633 | 3295 (1.6%) | 2028 (0.8%) |
| Anosmia | 0.5813 | <0.01 | [0.6 0.56] | 954.3881 | 7983 (3.8%) | 5525 (2.3%) |
| Taste sense altered | 0.6589 | <0.01 | [0.68 0.64] | 775.579 | 10426 (5.0%) | 8188 (3.3%) |
| Dyspnea | 0.8292 | <0.01 | [0.85 0.81] | 226.4865 | 13841 (6.6%) | 13601 (5.6%) |
| Chill | 0.8555 | <0.01 | [0.89 0.82] | 70.1969 | 5890 (2.8%) | 5928 (2.4%) |
| Fever | 0.8925 | <0.01 | [0.91 0.88] | 184.7625 | 32501 (15.5%) | 34562 (14.1%) |
| Sneezing | 0.9386 | <0.01 | [0.97 0.91] | 14.8189 | 7281 (3.5%) | 8024 (3.3%) |
| Vomiting | 0.9448 | <0.01 | [0.98 0.91] | 9.5393 | 5780 (2.8%) | 6408 (2.6%) |
| Nausea | 0.9719 | 0.2118 | [1.02 0.93] | 1.5588 | 3675 (1.8%) | 4187 (1.7%) |
| Malaise | 1.0145 | 0.6234 | [0.96 1.07] | 0.2411 | 2165 (1.0%) | 2573 (1.1%) |
| Sweating | 1.0382 | 0.1666 | [0.98 1.09] | 1.9136 | 2511 (1.2%) | 3053 (1.2%) |
| Fatigue | 1.0677 | <0.01 | [1.05 1.09] | 60.2058 | 29621 (14.2%) | 36704 (15.0%) |
| Chest pain | 1.0742 | <0.01 | [1.02 1.13] | 7.4219 | 2634 (1.3%) | 3312 (1.4%) |
| Limb pain | 1.1507 | <0.01 | [1.12 1.18] | 87.9483 | 8114 (3.9%) | 10876 (4.4%) |
| Headache | 1.1637 | <0.01 | [1.14 1.19] | 266.0105 | 22846 (10.9%) | 30601 (12.5%) |
| Dizziness | 1.1674 | <0.01 | [1.12 1.22] | 50.2551 | 3701 (1.8%) | 5047 (2.1%) |
| Cough | 1.2081 | <0.01 | [1.19 1.23] | 636.9268 | 38378 (18.4%) | 52325 (21.4%) |

| | | | | | | |
|---|---|---|---|---|---|---|
| Rhinorrhea | 1.3654 | <0.01 | [1.33 1.41] | 434.1302 | 7570 (3.6%) | 11952 (4.9%) |
| Nasal congestion | 1.4654 | <0.01 | [1.38 1.55] | 159.3566 | 1726 (0.8%) | 2952 (1.2%) |
| Concentration problems | 1.6352 | <0.01 | [1.58 1.7] | 673.4027 | 4285 (2.0%) | 8104 (3.3%) |
| Pain in throat | 1.9059 | <0.01 | [1.86 1.96] | 2327.0089 | 8381 (4.0%) | 18059 (7.4%) |
